# Impact and cost-effectiveness of nonavalent human papillomavirus vaccination in Switzerland: insights from a dynamic transmission model

**DOI:** 10.1101/19012674

**Authors:** Maurane Riesen, Johannes A. Bogaards, Nicola Low, Christian L. Althaus

## Abstract

**AIM:** In Switzerland, human papillomavirus (HPV) vaccination has been implemented using a quadrivalent vaccine that covers HPV types 16 and 18, responsible for about 70% of cervical cancer. The average national uptake was 56% in girls by the age of 16 years in 2014–2016. A nonavalent vaccine, covering five additional oncogenic HPV types was recommended at the end of 2018. The primary aim of this study was to assess the impact and cost-effectiveness of introducing the nonavalent HPV vaccine in Switzerland compared with the quadrivalent vaccine.

**METHODS:** We developed a dynamic transmission model that describes the spread of 10 high risk HPV types. We informed the model with Swiss data about sexual behaviour and cervical cancer screening, and calibrated the model to cervical cancer incidence in Switzerland. We modelled the impact of quadrivalent and nonavalent vaccines at the achieved (56%) and national recommended uptake (80%) in girls. We calculated the incremental cost-effectiveness ratio (ICER) between the nonavalent vaccine, the quadrivalent vaccine and no vaccination. We evaluated costs linked to cervical cancer screening, treatment of different disease stages and vaccination in a sensitivity analysis.

**RESULTS:** Compared with quadrivalent HPV vaccination in Switzerland at 56% uptake, vaccinating with the nonavalent vaccine would avert 1,175 cervical cancer deaths, 3,641 cases of cervical cancer and 106,898 CIN treatments over 100 years at 56% uptake. Compared with the quadrivalent vaccine, which would prevent an estimated 67% and 72% of cervical cancer cases at 56% and 80% coverage, the nonavalent vaccine would prevent 83% and 89% of all cervical cancers at the same coverage rates. The sensitivity analysis shows that introducing the nonavalent vaccination should improve health outcomes and offers a cost-saving alternative to the quadrivalent vaccine under the current price difference.

**CONCLUSIONS:** All scenarios with quadrivalent and nonavalent vaccination are likely to be cost-effective compared with no vaccination. Switching to the nonavalent vaccine at current and improved vaccination uptake is likely to be cost-saving under the investigated price difference.

## INTRODUCTION

Human papillomavirus (HPV) is the most common sexually transmitted virus [1]. HPVs cause cancer of the uterine cervix, the fourth most common cancer in women aged 15–44 years in Switzerland [2, 3]. Around 40 different HPV genotypes can infect anogenital squamous epithelium. These genotypes can be classified as high risk (HR) types, which can cause cervical and other anogenital cancers and low risk (LR) types, some of which cause anogenital warts. Vaccines containing synthetic virus-like particles are highly efficacious in the prevention of HPV infection and pre-cancerous lesions of the cervix (cervical intraepithelial neoplasia, CIN) [4]. As of 2019, there are three vaccines against HPV infections targeting two, four or nine HPV types. The bivalent vaccine (Cervarix^®^, GlaxoSmithKline, 2007) targets HPV 16 and 18, which are responsible for 70% of all cervical cancer cases. The quadrivalent vaccine (Gardasil^®^, Merck & Co., 2006) also targets two LR types (6, 11), which cause more than 90% of anogenital warts [5]. More recently, a nonavalent vaccine was developed (Gardasil-9^®^, Merck & Co.) targeting five HR types (31, 33, 45, 52, 58) in addition to those targeted by the quadrivalent vaccine, covering approximately 90% of all cervical cancers [6]. The nonavalent vaccine was first approved in the USA in 2014, followed by Canada, Australia and the European Union in 2015.

Mathematical modelling studies provide estimates of long-term impact and cost-effectiveness of HPV vaccination on cancer prevention and HPV elimination [7–13] and have supported the introduction of national vaccination strategies. Dynamic transmission models can take into account individual effects of vaccination on reducing the incidence of HPV infection over time, and herd effects, whereby vaccination of part of a population gives indirect protection to those who are unvaccinated. Various studies of the bivalent and quadrivalent HPV vaccines compared the HPV vaccination with cervical cancer screening only in different target groups and generally found that HPV vaccination is cost-effective [14–20]. Differences in results between studies reflect differences in model types, model assumptions, strategies assessed, disease outcomes included, settings, vaccine used and vaccination costs. The impact of introducing the nonavalent vaccine, which covers more HPV genotypes but is more expensive than the quadrivalent vaccine, has also been assessed in several countries and for different settings [6, 21–24]. To our knowledge, there are no cost-effectiveness studies of HPV vaccination that address the situation in Switzerland using a dynamic transmission model.

In Switzerland, the nonavalent vaccine was recommended by the Swiss Federal Office of Public Health (FOPH) and the Federal Commission for Vaccination in October 2018 [25, 26]. Until the end of 2018, 95% of vaccinated women had received the quadrivalent vaccine [27]. The national HPV vaccination strategy recommends: basic vaccination for girls aged 11–14 years (two doses at 0 and 6 months); catch-up vaccination for unvaccinated young women aged 15–19 (three doses at 0, 1 or 2, and 6 months); and complementary vaccination can be offered to unvaccinated women aged 20–26, boys aged 11–14 (two doses) and men aged 15–26 (three doses). HPV vaccination offered through a cantonal programme is free of charge for women aged 11–16 years and men aged 11–26 years [25]. Individual cantons are responsible for their own vaccination programmes. Some programmes are mainly based on school-based vaccination, others refer people to medical services such as paediatricians, general practitioners, gynaecologists, or hospitals. The FOPH estimates that national average for two-dose vaccination coverage in 16 years old girls was 56% (95% CI: 54.2%–58.7%) in the period 2014–2016, but uptake varies widely among cantons [28, 29]. The uptake rate in young men has not been reported to date.

The aim of this study was to assess the long-term impact and cost-effectiveness of introducing the nonavalent HPV vaccine in Switzerland compared with the quadrivalent vaccine. Since the national HPV vaccination strategy primarily targets girls, and estimates about HPV vaccine uptake in boys are not available at this date, we investigated girl-only vaccination strategies. To this end, we developed a mathematical model of HPV transmission that includes progression to pre-cancerous stages of cervical intraepithelial neoplasia (CIN) and to invasive cancer. We used Swiss data on sexual behaviour, screening and cervical cancer incidence to inform the model. We then performed a cost-effectiveness analysis to calculate the incremental cost-effectiveness ratio (ICER) between the nonavalent vaccine, the quadrivalent vaccine and no vaccination at current and intended vaccine uptake rates.

## METHODS

### HPV transmission model

We developed a compartmental transmission model for HPV that is formulated by ordinary differential equations (ODEs) and based on a previous model that we developed to investigate HPV transmission in Switzerland [30]. The susceptible-infected-recovered (SIR) structure was extended by adding stages for vaccination, CIN-2 and CIN-3, and three different cancer stages: localised, regional and distant cervical cancer that are either diagnosed or undiagnosed (fig. 1). We stratified the Swiss population into women and men, six age groups, and two sexual activity groups, and considered heterosexual mixing by age and sexual activity.

**Figure 1.**
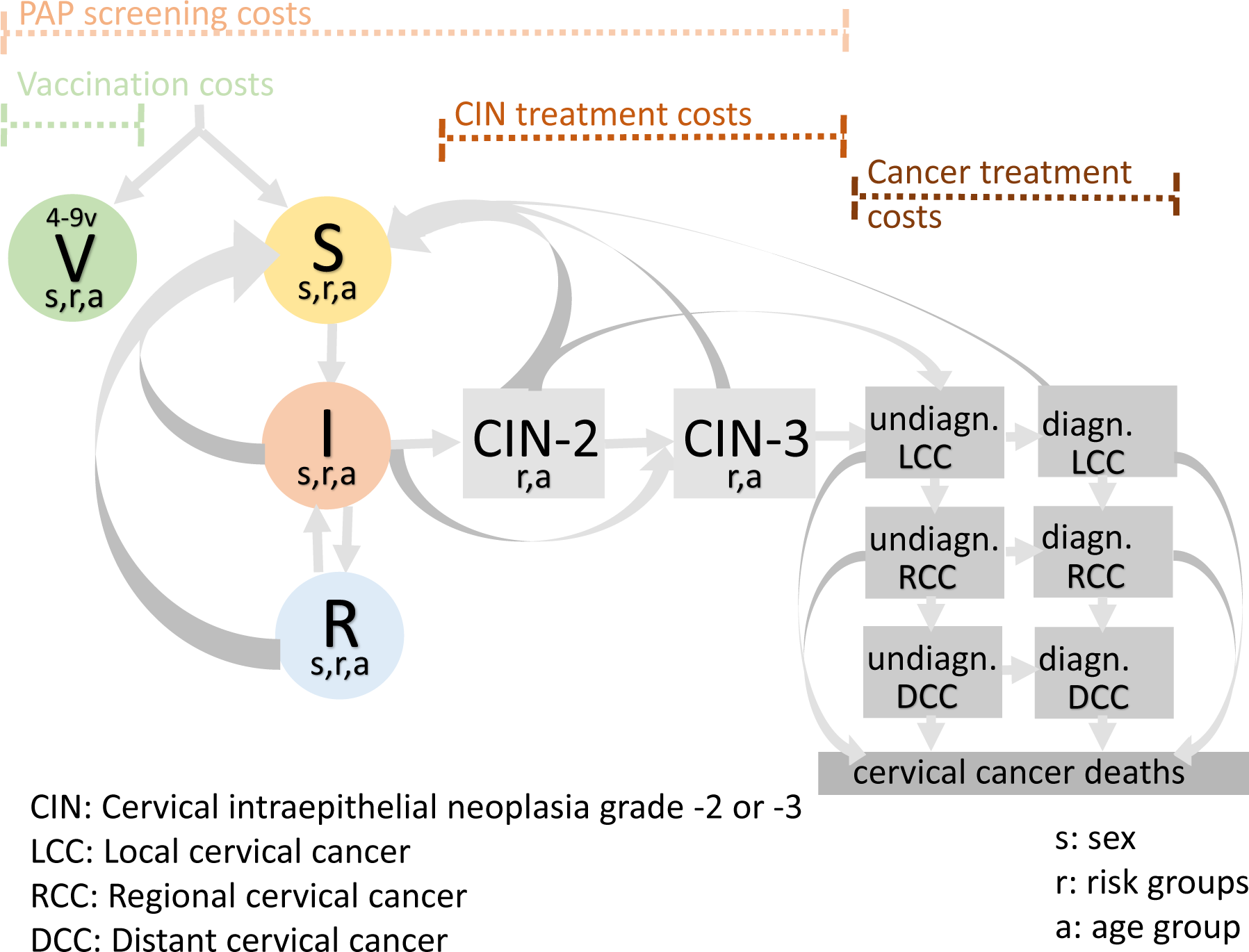
HPV transmission and disease progression model. The compartments for different infection and disease progression states are represented as circles and boxes, respectively. Individuals of sex *s*, age group *a* and sexual activity group *r* can either be vaccinated (*V*_*sra*_), susceptible (*S*_*sra*_), infected (*I*_*sra*_) or recovered (*R*_*sra*_). In women, HPV infection can progress to cervical intraepithelial neoplasia (CIN) of grade 2 or 3. Women who attend screening can be treated for CIN-2 and CIN-3, if timely detected. If left untreated, CIN-2 and CIN-3 can progress to three different cancer stages: localised, regional and distant cervical cancer that are either diagnosed or undiagnosed.

We modelled 10 different HR HPV types (16, 18, 31, 33, 35, 39, 45, 51, 52, 58) and considered all other HR types as one additional type. All types were modelled separately and were then grouped in a second step to assess the overall burden of HR HPV infections. Ignoring co-infections with multiple HPV types leads to an underestimation of the benefit of screening since lesions caused by multiple types in a single patient can be treated at the same time. Co-infection was accounted for by increasing the rate of cervical cancer screening by Papanicolau (Pap) test by the percentage of co-infections found in CIN3+ cases in Switzerland (12.8%) [31]. The transmission model and its calibration process is fully described in the appendix.

We integrated the ODEs until the system approached a steady-state for the pre-vaccination prevalence and incidence of HPV types, CIN stages and cervical cancer. We then simulated the different vaccination scenarios for 100 years. The model was run in the R software environment for statistical computing using the *deSolve* and *rootSolve* packages [32]. Model simulations were performed on UBELIX (http://www.id.unibe.ch/hpc), the high performance computing (HPC) cluster at the University of Bern. All code files can be downloaded from GitHub (https://github.com/mauraner/HPV_vacc_costeffect_Switzerland).

### Data and parameters

#### Age groups

We considered the following seven age groups: 10–14, 15–19, 20–24, 25–29, 30–39, 40–64, and 65–99 year olds. For simplicity, we ignored background mortality and changes in demography over time, which results in a uniform age distribution in the population. However, we weighted the number of individuals in each age group according to the Swiss population structure in 2016 and a total population of 8.48 million (appendix, table A1) [33].

#### Sexual behaviour

We used data from the Swiss Health Survey (SHS) to inform sexual behaviour [34]. The SHS has a limited number of questions about sexual behaviour, which do not permit the calculation of the rate of sexual partner change. To do this, we used data that are available from the third British National Survey of Sexual Attitudes and Lifestyles (Natsal-3), a national cross-sectional survey of 15,162 women and men aged 16–74 years conducted in 2010–2012 [35–37]. We calculated the sex- and age-stratified probability that an individual with *x* total sexual partners had *y* new heterosexual partners in the last year (appendix, section 1.5 and fig. A1). The first age group (10–14 years) was assumed to be sexually inactive. Sexual mixing between age groups was also informed by Natsal-3 data.

#### HPV prevalence

To our knowledge there are no data about HR HPV prevalence that are representative of young women in Switzerland. We used data about HPV prevalence measured in urine samples collected from a random sub-sample of the Natsal-3 survey population to calibrate the model to HR HPV prevalence [37]. To account for later sexual initiation and fewer number of sexual partnerships for the Swiss compared to the British population [30], we assumed that HPV prevalence among Swiss women aged 20–24 years was comparable to that of 15–19 year old women in Natsal-3. We calibrated the model to 24.4% HR HPV prevalence in 20–24 year old women. This point estimate of HR HPV prevalence is situated within the confidence interval of an estimate from women aged 21–25 years who were taking part in a multi-centre diagnostic test evaluation study of liquid-based cytology and were considered at low risk of cervical cancer [38]. Individual contributions of different HR HPV types to the overall prevalence were informed by data from a systematic review, which included 423 studies worldwide that measured the presence of HPV using molecular diagnostic methods (appendix, fig. A4 and table A6) [39].

#### Cancer incidence

We used cervical cancer incidence data from the Swiss National Institute for Cancer Epidemiology and Registration (NICER) [40]. We calibrated the pre-vaccination steady-state of our model to the crude incidence from 2004–2014 (6.4 cases per 100,000 women, respectively 7.2 cases for the age groups included in the model). To attribute cervical cancer cases to different HPV types, we used data from a meta-analysis presenting results of 2,058 cases of invasive cervical cancer in Europe (excluding 34 cases of low-risk types 6, 11, 42, 53, 66, and 73) [41]. For simplicity, we assumed that 100% of the cervical cancer cases were caused by HPV infection [41, 42]. We scaled the modelled incidence of cervical cancer to a Swiss population of 8.48 million, assuming a sex ratio of 1:1 [43].

#### Vaccination

We only consider the vaccinated population targeted by the basic vaccination strategy (11–14 year old girls) and whose uptake rates are reported in the Swiss National Vaccination Coverage Survey [44]. Therefore, the girls enter the system being vaccinated and the protective effect of vaccination starts having an impact on the first sexually active age group (15–19 year olds). We do not take into account the cantonal differences in vaccination uptake.

#### Cervical cancer screening

Cervical cancer screening rates were derived from the SHS, which contains information on whether women have had a Pap smear in the last 12 months or ever (7,881 and 10,124, respectively) (appendix, fig. A2). We calculated the proportion of women who have had a Pap smear in the last 12 months for each age group. We did not have data for 15–19 year olds and assumed that cervical screening started from age 20 onwards. For the model, we transformed the proportion of women with a screen to yearly rates (appendix, table A4). The screening test sensitivity and specificity for CIN-2 or CIN-3 were 59% and 96%, respectively [20, 38].

### Cost-effectiveness analysis

The cost-effectiveness analysis was conducted from a healthcare system perspective. We considered the costs of Pap smears, colposcopy in case of abnormal cytology, treatment of CIN-2 and CIN-3, treatment of different cancer stages and vaccination costs (table 1). We assumed the cost per single dose of the quadrivalent vaccine to be CHF 90.30 (66.60 for the vaccine dose and CHF 23.70 for the medical procedure) [45]. The per-dose factory price difference between the quadrivalent and nonavalent vaccine for the private market in Switzerland was CHF 4.16 (personal communication with MSD Merck Sharp & Dohme AG). We used the costs of screening from a previous cost-effectiveness study for Switzerland [20]. Costs for different cervical cancer stages were calculated by using CHF 20,000 as a baseline cost for cervical cancer treatment [20] and adding the cost difference for different cancer stages assuming the cost differences indicated in Durham et al. [20, 21]. We based the health utility indices for different disease stages on previous literature [19, 21, 46–49]. Costs and quality-adjusted life years (QALYs) are further described in the appendix (table A5). The cost-effectiveness analysis was calculated for a time period of 100 years after vaccination onset using a 3% annual discounting rate. This time horizon is consistent with the recommendations of the World Health Organization, European Vaccine Economics Community for dynamic models and with other HPV vaccination cost-effectiveness studies [24, 50–53].

**Table 1.** Distribution and range of cost-parameters from the multivariable sensitivity analysis.

| Parameter | Baseline value | Range* | Distribution |
| --- | --- | --- | --- |
| Discounting rate | 3% | 0-6% | uniform |
| CIN-2 treatment costs (CHF) | 1,151 | 727–1,401 | log-normal |
| CIN-3 treatment costs (CHF) | 2,238 | 1,463–2,699 | log-normal |
| Local cancer treatment costs (CHF) | 20,000 | 13,426–20,312 | log-normal |
| Regional cancer treatment costs (CHF) | 21,890 | 14,426–26,945 | log-normal |
| Distant cancer treatment costs (CHF) | 39,000 | 25,453–46,967 | log-normal |
| Cost per negative screen (CHF) | 210 | 139–253 | log-normal |
| Cost per false-positive screen (CHF) | 80 | 51–100 | log-normal |
| Vaccination price difference, two doses (CHF) | 8.32 | 0.83–83.20 | uniform |
| V4v vaccination price (CHF) | 181 | 155–203 | log-normal |
| Utility score for CIN-2 | 0.92 | 0.87–0.97 | uniform |
| Utility score for CIN-3 | 0.91 | 0.86–0.96 | uniform |
| Utility score for local cancer | 0.76 | 0.68–0.84 | uniform |
| Utility score for regional cancer | 0.67 | 0.60–0.74 | uniform |
| Utility score for distant cancer | 0.48 | 0.42–0.53 | uniform |

\*Total range for uniform distribution and interquartile range for log-normal distribution
CIN-2/3, cervical intraepithelial neoplasia grade 2 or 3; CHF, Swiss francs; V4v, quadrivalent vaccine

We set the cost-effectiveness threshold to CHF 79,104 per QALY, which corresponds to the Swiss per capita gross domestic product (GDP) in 2017 [50, 54, 55]. We plotted the results from the sensitivity analysis on a cost-effectiveness plane, reporting the incremental QALY gained and the incremental costs in millions of CHF. We also presented the ICER as follows:

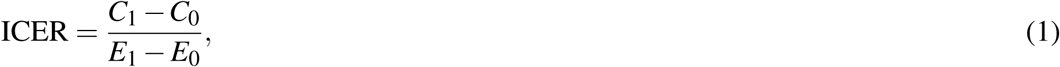

where *C*_1_ and *E*_1_ represent the costs and health effects of the desired intervention and *C*_0_ and *E*_0_ represent the costs and health effects of the comparator. The desired interventions are quadrivalent and nonavalent vaccination at either 56% or 80% uptake in girls and the comparators are no vaccination at all or quadrivalent vaccination at the same levels of vaccination coverage.

### Sensitivity analysis

The sensitivity analyses were performed in two steps: first 31 parameters influencing the health outcome were varied using Latin hypercube sampling of uniformed distributed values of the confidence intervals around the parameters (Supplementary material, table S4). We simulated 2,000 parameters sets for the 11 different HPV types and excluded 884 simulations which did not match the Swiss cervical cancer incidence or HPV prevalence. In a second step, with the other 1,116 simulations, we performed a sensitivity analysis on the parameters used for the cost-effectiveness assessment. These were varied around the baseline estimates using random uniform (discounting, QALYs weight, cost difference) and lognormal (quadrivalent vaccine cost, treatment costs) distributions (table 1). The shape parameters of the lognormal distributions were defined by assuming a standard deviation being half of the original baseline value. The range of the uniform distribution was defined as a ten-fold increase/decrease of the price difference between the two vaccines, and a 5% or 10% increase/decrease of the utility scores (for precancerous and cancerous stages, respectively). We also performed a univariable sensitivity analysis of the ICER with respect to changes in disease treatment costs, vaccination costs, the vaccination price difference and the discounting rate [50]. To this end, we used the mean and the maximum and minimum values from parameters varied uniformly (discounting, QALYs weight, cost difference) and interquartile range of parameters varied within a lognormal distribution (vaccination cost, treatment cost). We then showed the median ICERs for the mean, minimum, and maximum range of variation of each parameter assessed in the univariable analysis. The discounting rate was only varied in the univariable sensitivity analysis; in the multivariable analysis the baseline value of 3% was used for all parameter sets.

## RESULTS

### Impact of HPV vaccination

The results from the HPV transmission model, before vaccination onset, were in good agreement with empirical data. First, our model calibration resulted in pre-vaccination HR HPV prevalences per age-group that were in accordance with HR HPV positivity from a Swiss multi-centre diagnostic test evaluation study of liquid-based cytology [38]. Second, we obtained a very similar HPV type ratio in CIN-3 cases when compared to data from a Swiss study [31] and a meta-analysis for European populations [39] (appendix, fig. A4). Third, our model estimates (median, IQR) of 4,461 (3,798–5,082) CIN-2 and CIN-3 diagnoses and 99 (84-113) yearly deaths from cervical cancer during the pre-vaccination era were in line with previous reports of 5,000 CIN diagnoses and 90 deaths [56].

We modelled the effects of HPV vaccination on the number of cervical cancer cases, CIN diagnoses and cervical cancer death (fig. 2). After 100 years of HPV vaccination, the yearly age-standardised cervical cancer incidence decreased from 6.4 to 2.1 (1.9–2.3) per 100,000 women under the baseline vaccination strategy (56% quadrivalent vaccination uptake in young women). This corresponds to a reduction of 67.0% (64.4–70.4). The modelled incidence decreased by 72.4% using the quadrivalent vaccine at an uptake of 80%. Using the nonavalent vaccine, the modelled incidence decreased by 83.1% (80.3–86.6) at 56% uptake and by 88.6% at 80% uptake. Scenarios with 80% vaccination uptake are above the vaccination threshold resulting from the model. Therefore, no IQR is available for these scenarios since, in our simulations, elimination of vaccine-targeted HPV types is reached after 100 years at this level of coverage. The model estimates that, compared with 56% quadrivalent vaccination, the switch to nonavalent vaccination would incrementally avert 106,898 CIN treatments, 3,641 cervical cancer cases and 1,175 cervical cancer deaths over a time period of 100 years (table 2).

**Table 2.** Number of averted CIN cases (diagnosed), cervical cancer (CC) cases, and deaths over 100 years (median, IQR) for each vaccination strategy compared with no vaccination (upper part) and for nonavalent compared to quadrivalent vaccine given same coverage rates (lower part, *V*4*v*_56_, *V*4*v*_80_, *V*9*v*_56_, and *V*9*v*_80_ correspond to quadrivalent and nonavalent vaccination at an uptake of 56% and 80%, respectively). The number of cases are based on an age-standardised Swiss population of 8.48 million assuming a sex ratio of 1:1.

| Scenario | CIN diagnosed cases | CC cases | CC deaths |
| --- | --- | --- | --- |
| Averted cases with all vaccination strategies compared to no vaccination |  |  |  |
| V4v<sub>56</sub> | 195,164 (166,569-222,123) | 14,025 (13,571-14,431) | 4,443 (3,724-5,137) |
| V4v<sub>80</sub> | 225,287 (192,977-255,909) | 16,117 (16,015-16,217) | 5,148 (4,356-5,970) |
| V9v<sub>56</sub> | 302,196 (256,880-344,167) | 17,661 (17,218-18,084) | 5,619 (4,717-6,494) |
| V9v<sub>80</sub> | 335,219 (284,903-382,418) | 19,867 (19,747-19,979) | 6,359 (5,388-7,379) |
| Averted cases with nonavalent compared to quadrivalent vaccine |  |  |  |
| V9v<sub>56</sub> | 106,898 (89,933-122,885) | 3,641 (3,615-3,665) | 1,175 (992-1,357) |
| V9v<sub>80</sub> | 110,317 (92,847-126,702) | 3,751 (3,728-3,770) | 1,214 (1,027-1,399) |
| V4v<sub>56</sub>, quadrivalent vaccine and 56% uptake; V4v<sub>80</sub>, quadrivalent vaccine and 80% uptake; V9v<sub>56</sub>, nonavalent vaccine and 56% uptake; V9v<sub>80</sub>, nonavalent vaccine and 80% uptake |  |  |  |

**Figure 2.**
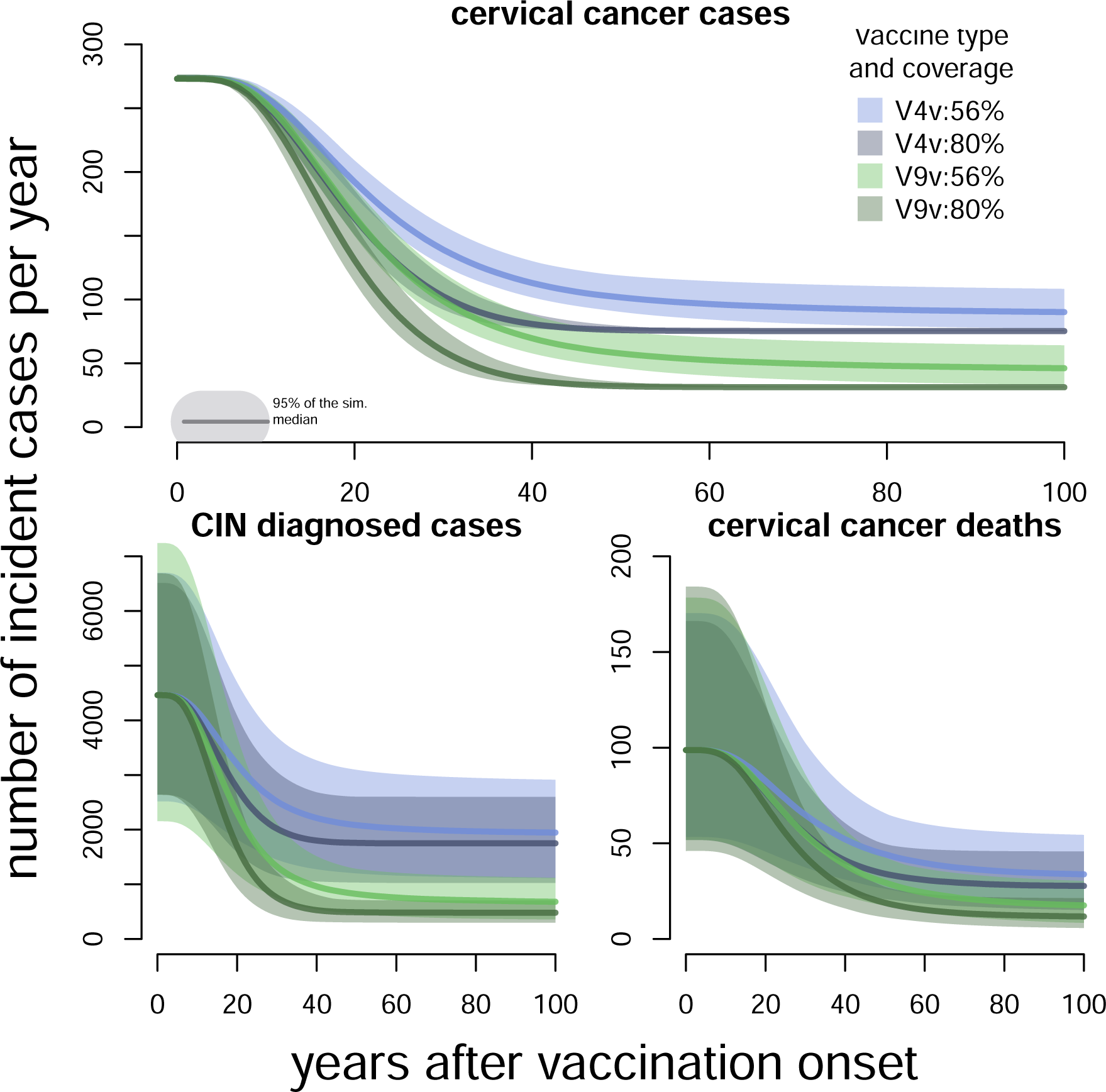
Reduction in CIN diagnoses, cervical cancer cases and mortality after introducing HPV vaccination. V4v (blue lines) and V9v (green lines) correspond to quadrivalent and nonavalent HPV vaccination, respectively. The number of cases are based on an age-standardised Swiss population of 8.48 million assuming a sex ratio of 1:1.

### Cost-effectiveness analysis

Compared with no vaccination, all vaccination scenarios are below the cost-effectiveness threshold corresponding to the Swiss GDP (fig. 3 and 4, left panels). The nonavalent HPV vaccine is likely to be cost-saving compared with the quadrivalent vaccine at the same vaccination coverage (fig. 3, right panel). For 56% vaccination coverage, we calculated an ICER (median, IQR) of −2,125 (−4,003 – −189) CHF per QALY (fig. 4). In both univariate and multivariate sensitivity analyses, the ICER remained negative over the whole range of the vaccination price difference for two doses (CHF 0.83 – 83.20). If the vaccination increased to 80%, nonavalent vaccine is also likely to be cost-saving compared to quadrivalent with an ICER of - 1,184 (−3,344 – 1,227). Variation in the discounting rate and higher vaccination price differences can lead to positive ICERs. Furthermore, the ICER is slightly higher and thus less cost-saving at 80% compared to 56% uptake. This is because the benefit of herd effects is more pronounced at 56% than at 80% coverage since 80% uptake exceeds the vaccination threshold of the model.

**Figure 3.**
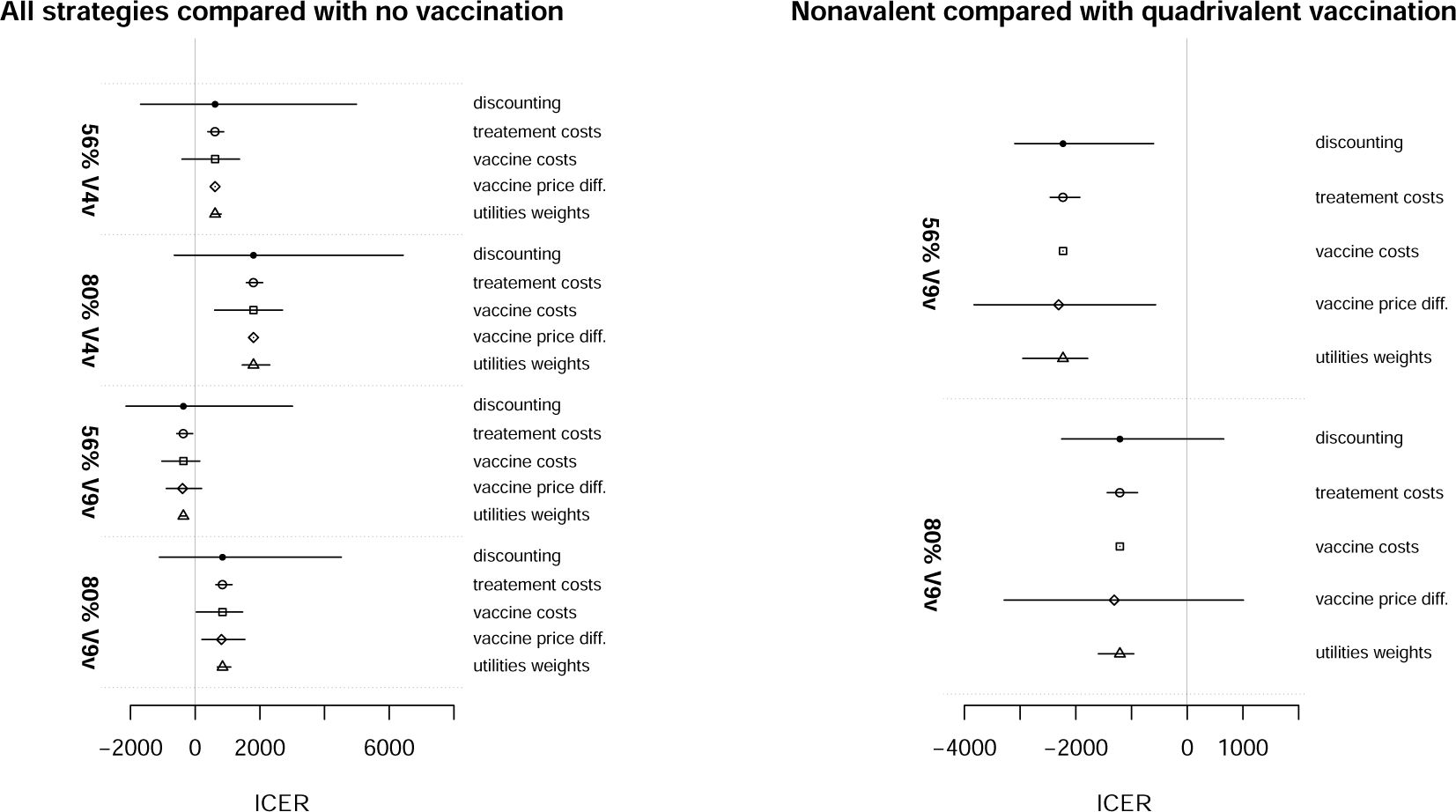
Univariate sensitivity analysis of ICER (median, IQR) comparing all vaccination strategies with no HPV vaccination (left panel) and quadrivalent vaccination with nonavalent given the same coverages (right panel). *V*4*v*_56_ means quadrivalent vaccine and 56% uptake, *V*4*v*_80_ quadrivalent vaccine and 80% uptake, *V*9*v*_56_ nonavalent vaccine and 56% uptake, *V*9*v*_80_ nonavalent vaccine and 80% uptake.

**Figure 4.**
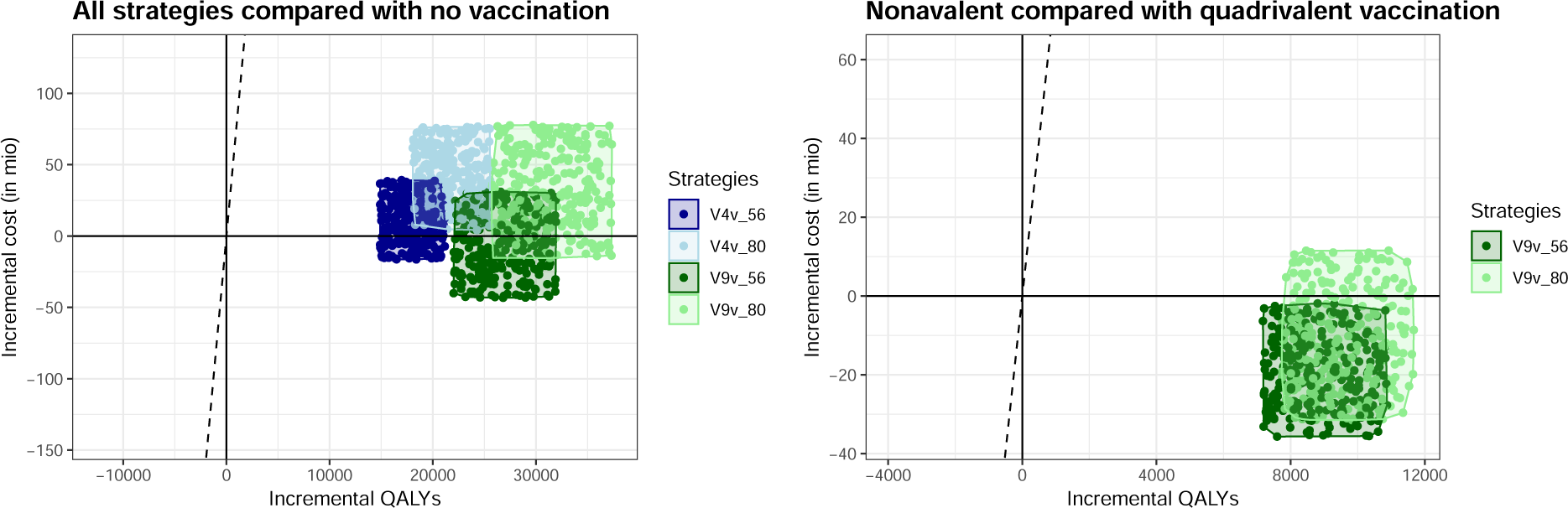
Multivariate cost-effectiveness planes comparing all vaccination strategies with no HPV vaccination (left panel) and quadrivalent vaccination with nonavalent given the same coverages (right panel). The dots represent simulations situated within the IQR of incremental cost and QALY values for each vaccination strategy. *V*4*v*_56_ means quadrivalent vaccine and 56% uptake, *V*4*v*_80_ quadrivalent vaccine and 80% uptake, *V*9*v*_56_ nonavalent vaccine and 56% uptake, *V* 9*v*_80_ nonavalent vaccine and 80% uptake.

## DISCUSSION

We developed a dynamic HPV transmission model, which we used to study the impact and cost-effectiveness of HPV vaccination of adolescent girls in Switzerland. Switching from the quadrivalent to the nonavalent vaccine at a vaccination uptake of 56% is expected to avert more than 100,000 CIN treatments, 3,000 cervical cancer cases, and 1,000 deaths over a time period of 100 years. All considered vaccination scenarios (quadrivalent or nonavalent at 56% or 80% uptake) are likely to be cost-effective given a cost-effectiveness threshold corresponding to the Swiss GDP. Switching from the quadrivalent to the nonavalent vaccine is likely to be cost-saving under the investigated price difference between the two vaccines.

To our knowledge, this is the first cost-effectiveness study of HPV vaccination in Switzerland using a dynamic transmission model. The model incorporates important features, such as multiple HR HPV genotypes and heterogeneity in sexual activity. We used Swiss data for sexual behaviour, cervical cancer screening and cervical cancer incidence. Calibrating the model to HPV prevalence and cervical cancer incidence resulted in CIN cases and deaths from cervical cancer that were in good agreement with empirical data.

This study comes with a number of limitations. First, there were no population-representative data about HPV prevalence in young Swiss women available for the model calibration. Since our previous modelling study identified differences in sexual behaviour between the Swiss and British populations [30], we used a simplified assumption that the HR HPV prevalence in Swiss 20–24 year-old women could be compared with a younger age group from the Natsal-3 study. Despite this, the resulting HPV prevalences in older age group and the resulting number of diagnosed CIN cases per year were in accordance with available data from Swiss studies [38, 56].

Second, we assumed that all diagnosed CIN-2 or CIN-3 cases were treated. In practice, this might not be the case if some women are lost to follow-up, or receive active monitoring rather than immediate treatment. Therefore, the number of treated cases estimated by the model is higher than estimates of treated CIN cases in Switzerland, although the number of diagnoses corresponds [56]. However, given the range of treatment costs included in the sensitivity analysis, we do not expect that this difference would have an important effect on the estimated ICERs. Third, we used health utility scores using published values from the literature with little information on their uncertainty. Our sensitivity analysis showed that the cost-effectiveness analysis can be influenced by small variations in these estimates in some scenarios. Fourth, we simulated the different vaccination scenarios from the same pre-vaccination steady-state and did not explicitly take into account that a period of quadrivalent vaccination precedes the implementation of nonavalent vaccination. Given that the differences between scenarios are rather small during the first years of vaccination, we do not expect this simplification to have a considerable impact on our results. Finally, we did not include catch-up vaccination in women or complementary vaccination in boys because we had no data on vaccination uptake in these sub-populations. Including catch-up vaccination and vaccination in boys would likely result in an increase of the ICERs when compared to no vaccination [10, 15, 18, 57].

Our results about the general health benefits of introduction the nonavalent vaccine are in line with similar studies for other high-income countries [21, 22, 24, 58–61]. In contrast, we found that the nonavalent HPV vaccine averts a much higher number of episodes of CIN-2 and CIN-3 than two other modelling studies from Switzerland and Austria [24, 62]. The authors of the Austrian study acknowledged that their model substantially underestimated CIN incidence so our model probably better describes this aspect. Despite this difference, our study is in agreement with the results from the Austrian study that vaccination strategies with the nonavalent vaccine are cost-effective compared to the quadrivalent vaccination strategies. Our study further corroborates previous findings suggesting that nonavalent HPV vaccination is likely to be cost-effective in Switzerland [20]. Our study has advantages over the only other published cost-effectiveness study of HPV vaccination in Switzerland, which used a static Markov model [20]. Our dynamic transmission model accounts for the indirect herd effects of vaccination, which is a crucial aspect in vaccination against infectious diseases [63, 64].

The cost-effectiveness of nonavalent HPV vaccine in women depends on the balance between the additional reduction in CIN and invasive cancer because of the inclusion of more HR HPV genotypes and the cost difference. In Switzerland, the increase in the price for two doses of the nonvalent HPV vaccine, compared to the quadrivalent vaccine, is small (CHF 8.32). Nevertheless, our sensitivity analysis shows that the nonavalent HPV vaccine would be cost-saving over a ten-fold increase in the price difference (CHF 83.20) at the current uptake of 56%. Drolet et al. [22] found that the additional cost per dose of the nonavelent vaccine should not exceed $11 (vaccinating girls at 80% coverage) in order to remain cost-saving compared to the quadrivalent vaccine. This is more than twice the baseline per-dose price difference that we used for our study. ICERs for our modelled vaccination strategies compared with no vaccination were generally lower than estimates from other studies in similar settings [20, 22, 52]. This difference can be partly explained by the two-dose vaccination schedule in Switzerland, which is cheaper than a three-dose schedule [65–67]. Furthermore, many of the other studies included scenarios with male vaccination which can lead to higher ICERs.

Future modelling studies could include disease outcomes other than cervical cancer, such as genital warts or anal, oropharyngeal, penile, vaginal, and vulvar cancers [68, 69]. In high-income countries, non-cervical cancers represent one quarter of all cancers caused by HPV [70]. In Switzerland, we currently do not have precise estimates of the incidence of these additional outcomes. We expect that adding the effect of vaccination on genital warts and non-cervical cancers would further decrease the ICER of the four different vaccination scenarios compared to the situation without vaccination. However, it would not affect the comparison between the quadrivalent and the nonavalent vaccine.

Further research and cost-effectiveness analyses are needed to assess the effects of male vaccination and the impact of vaccination on cancers other than cervical cancer [71–73]. For informative modelling studies, additional empirical data about the uptake of male HPV vaccine and the HPV-attributable fraction and natural history of cancers such as squamous cancer of the oropharynx are, however, essential. Further studies should also investigate optimisation of cervical cancer screening recommendations (time interval, screening method) in the light of nonavalent vaccine use and of different vaccination uptake scenarios. It is likely that reduced number of cervical screens per life time and a change to HPV genotyping as primary screening method would be better with a switch to nonavalent vaccination and increased uptake rates [74, 75]. The Swiss Society of Gynaecology and Obstetrics has recently revised its screening guidelines and now recommends the use of HPV test as first screening method in women aged 30 or more [76]. The use of HPV test as primary triage method might be worth extending to all age groups in vaccinated women or in all women if the vaccination uptake increases [77]. The results of this study suggest that the introduction of the nonavalent HPV vaccine was likely to be cost-saving compared to the quadrivalent vaccine at the current and improved vaccination uptake in Switzerland under current pharmacy price difference.

## Data Availability

All code files can be downloaded from GitHub: https://github.com/mauraner/HPV_vacc_costeffect_Switzerland.

## REFERENCES

1 Centers for Disease Control and Prevention (CDC). STD Facts - Human papillomavirus (HPV); 2017. Available from: https://www.cdc.gov/std/hpv/stdfact-hpv.htm.

2 Schiffman M, Castle PE, Jeronimo J, Rodriguez AC, Wacholder S. Human papillomavirus and cervical cancer. Lancet (London, England). 2007 Sep;370(9590):890–907.

3 Bruni L, Barrionuevo-Rosas L, Alberro G, Serrano B, Mena M, Gomez D, et al. Human Papillomavirus and Related Diseases Report in Switzerland; 2017. Available from: www.hpvcentre.net.

4 Harper DM, Franco EL, Wheeler C, Ferris DG, Jenkins D, Schuind A, et al. Efficacy of a bivalent L1 virus-like particle vaccine in prevention of infection with human papillomavirus types 16 and 18 in young women: a randomised controlled trial. Lancet. 2004;364(9447):1757–65.

5 Giuliano AR, Tortolero-Luna G, Ferrer E, Burchell AN, de Sanjose S, Kjaer SK, et al. Epidemiology of Human Papillomavirus Infection in Men, Cancers other than Cervical and Benign Conditions. Vaccine. 2008 Aug;26:K17–K28. Available from: http://www.sciencedirect.com/science/article/pii/S0264410X08007469.

6 Hartwig S, Baldauf JJ, Dominiak-Felden G, Simondon F, Alemany L, de Sanjosé S, et al. Estimation of the epidemiological burden of HPV-related anogenital cancers, precancerous lesions, and genital warts in women and men in Europe: Potential additional benefit of a nine-valent second generation HPV vaccine compared to first generation HPV vaccines. Papillomavirus Research. 2015 Dec;1:90–100. Available from: http://www.sciencedirect.com/science/article/pii/S2405852115000099.

7 Pink J, Parker B, Petrou S. Cost Effectiveness of HPV Vaccination: A Systematic Review of Modelling Approaches. PharmacoEconomics. 2016 Sep;34(9):847–861. Available from: http://link.springer.com/10.1007/s40273-016-0407-y.

8 Suijkerbuijk AWM, Donken R, Lugnér AK, de Wit GA, Meijer CJLM, de Melker HE, et al. The whole story: a systematic review of economic evaluations of HPV vaccination including non-cervical HPV-associated diseases. Expert Review of Vaccines. 2017;16(4):361–375.

9 Favato G, Easton T, Vecchiato R, Noikokyris E. Ecological validity of cost-effectiveness models of universal HPV vaccination: A systematic literature review. Vaccine. 2017 May;35(20):2622–2632. WOS:000401214100003.

10 Ng SS, Hutubessy R, Chaiyakunapruk N. Systematic review of cost-effectiveness studies of human papillomavirus (HPV) vaccination: 9-Valent vaccine, gender-neutral and multiple age cohort vaccination. Vaccine. 2018;36(19):2529–2544.

11 Gervais F, Dunton K, Jiang Y, Largeron N. Systematic review of cost-effectiveness analyses for combinations of prevention strategies against human papillomavirus (HPV) infection: a general trend. BMC Public Health. 2017 Mar;17(1):283. Available from: https://doi.org/10.1186/s12889-017-4076-3.

12 Brisson M, Edmunds WJ. Economic evaluation of vaccination programs: the impact of herd-immunity. Med Decis Making. 2003;23(1):76–82.

13 Brisson M, Edmunds WJ. Impact of model, methodological, and parameter uncertainty in the economic analysis of vaccination programs. Med Decis Making. 2006;26(5):434–46.

14 Brisson M, Bénard É, Drolet M, Bogaards JA, Baussano I, Vänskä S, et al. Population-level impact, herd immunity, and elimination after human papillomavirus vaccination: a systematic review and meta-analysis of predictions from transmission-dynamic models. The Lancet Public Health. 2016 Nov;1(1):e8–e17. Available from: http://www.sciencedirect.com/science/article/pii/S2468266716300019.

15 Bogaards JA, Kretzschmar M, Xiridou M, Meijer CJLM, Berkhof J, Wallinga J. Sex-specific immunization for sexually transmitted infections such as human papillomavirus: insights from mathematical models. PLoS medicine. 2011 Dec;8(12):e1001147.

16 Jit M, Brisson M, Portnoy A, Hutubessy R. Cost-effectiveness of female human papillomavirus vaccination in 179 countries: a PRIME modelling study. The Lancet Global Health. 2014 Jul;2(7):e406–e414. Available from: http://www.sciencedirect.com/science/article/pii/S2214109X14702372.

17 Goldie SJ, Kim JJ, Wright TC. Cost-effectiveness of human papillomavirus DNA testing for cervical cancer screening in women aged 30 years or more. Obstetrics and Gynecology. 2004 Apr;103(4):619–631.

18 Kim JJ, Goldie SJ. Cost effectiveness analysis of including boys in a human papillomavirus vaccination programme in the United States. BMJ. 2009 Oct;339:b3884. Available from: https://www.bmj.com/content/339/bmj.b3884.

19 Chesson HW, Ekwueme DU, Saraiya M, Dunne EF, Markowitz LE. The cost-effectiveness of male HPV vaccination in the United States. Vaccine. 2011 Oct;29(46):8443–8450. Available from: http://linkinghub.elsevier.com/retrieve/pii/S0264410X11011467.

20 Szucs TD, Largeron N, Dedes KJ, Rafia R, Bénard S. Cost-effectiveness analysis of adding a quadrivalent HPV vaccine to the cervical cancer screening programme in Switzerland. Current Medical Research and Opinion. 2008 May;24(5):1473–1483. Available from: https://www.tandfonline.com/doi/full/10.1185/030079908X297826.

21 Durham DP, Ndeffo-Mbah ML, Skrip LA, Jones FK, Bauch CT, Galvani AP. National- and state-level impact and cost-effectiveness of nonavalent HPV vaccination in the United States. Proceedings of the National Academy of Sciences of the United States of America. 2016 May;113(18):5107–5112.

22 Drolet M, Laprise JF, Boily MC, Franco EL, Brisson M. Potential cost-effectiveness of the nonavalent human papillomavirus (HPV) vaccine. International Journal of Cancer. 2014;134(9):2264–2268. Available from: https://onlinelibrary.wiley.com/doi/abs/10.1002/ijc.28541.

23 Kiatpongsan S, Kim JJ. Costs and Cost-Effectiveness of 9-Valent Human Papillomavirus (HPV) Vaccination in Two East African Countries. PLOS ONE. 2014 Sep;9(9):e106836. Available from: https://journals.plos.org/plosone/article?id=10.1371/journal.pone.0106836.

24 Boiron L, Joura E, Largeron N, Prager B, Uhart M. Estimating the cost-effectiveness profile of a universal vaccination programme with a nine-valent HPV vaccine in Austria. BMC Infectious Diseases. 2016 Apr;16. Available from: https://www.ncbi.nlm.nih.gov/pmc/articles/PMC4833954/.

25 Spaar A, Heininger U, Stronski Huwiler S, Masserey Spicher V. Die HPV-Impfung ist wirksam und sicher. BAG-Bulletin. 2018 Jan;Woche 3/2018:9.

26 Federal Office of Public Health. HPV-Impfung: Empfehlungen des BAG und der EKIF zum neuen Impfstoff Gardasil 9 R. 2018 Oct;43:10–15.

27 Federal Office of Public Health. Die HPV-Impfung in der Schweiz: Resultate einer nationalen Befragung im Jahr 2014. BAG-Bulletin. 2015 Jun;23/15.

28 Riesen M, Konstantinoudis G, Lang P, Low N, Hatz C, Maeusezahl M, et al. Exploring variation in human papillomavirus vaccination uptake in Switzerland: a multilevel spatial analysis of a national vaccination coverage survey. BMJ Open. 2018 May;8(5):e021006. Available from: https://bmjopen.bmj.com/content/8/5/e021006.

29 Federal Office of Public Health. Durchimpfung von 2-,8-und 16-Jährigen in der Schweiz, 2011 bis 2013. Bulletin. 2015 Jul;28.

30 Riesen M, Garcia V, Low N, Althaus CL. Modeling the consequences of regional heterogeneity in human papillomavirus (HPV) vaccination uptake on transmission in Switzerland. Vaccine. 2017 Dec;35(52):7312–7321. Available from: http://www.sciencedirect.com/science/article/pii/S0264410X17315323.

31 Egli-Gany D, Spaar Zographos A, Diebold J, Masserey Spicher V, Frey Tirri B, Heusser R, et al. Human papillomavirus genotype distribution and socio-behavioural characteristics in women with cervical pre-cancer and cancer at the start of a human papillomavirus vaccination programme: the CIN3plus-plus study. BMC cancer. 2019 Jan;19(1):111.

32 R Core Team. R: A Language and Environment for Statistical Computing. Vienna, Austria; 2016.

33 Federal Statistical Office. Statistique de la population et des ménages STATPOP - Population résidante permanente et non permanente. Neuchâtel, Suisse; 2017. Available from: www.bfs.admin.ch.

34 Federal Statistical Office. Swiss Health Survey / Schweizerische Gesundheitsbefragung / Enquête suisse sur la santé / Indagine sulla salute in Svizzera. Neuchâtel, Suisse; 2012. Available from: https://www.bfs.admin.ch/bfs/de/home/statistiken/gesundheit/erhebungen/sgb.html.

35 Mercer CH, Tanton C, Prah P, Erens B, Sonnenberg P, Clifton S, et al. Changes in sexual attitudes and lifestyles in Britain through the life course and over time: findings from the National Surveys of Sexual Attitudes and Lifestyles (Natsal). The Lancet. 2013 Nov;382(9907):1781–1794. Available from: http://www.sciencedirect.com/science/article/pii/S0140673613620358.

36 Sonnenberg P, Clifton S, Beddows S, Field N, Soldan K, Tanton C, et al. Prevalence, risk factors, and uptake of interventions for sexually transmitted infections in Britain: findings from the National Surveys of Sexual Attitudes and Lifestyles (Natsal). The Lancet. 2013 Nov;382(9907):1795–1806. Available from: http://www.sciencedirect.com/science/article/pii/S0140673613619479.

37 Erens B, Phelps A, Clifton S, Mercer CH, Tanton C, Hussey D, et al. Methodology of the third British National Survey of Sexual Attitudes and Lifestyles (Natsal-3). Sexually Transmitted Infections. 2014 Mar;90(2):84–89. Available from: http://sti.bmj.com/lookup/doi/10.1136/sextrans-2013-051359.

38 Bigras G, de Marval F. The probability for a Pap test to be abnormal is directly proportional to HPV viral load: results from a Swiss study comparing HPV testing and liquid-based cytology to detect cervical cancer precursors in 13 842 women. British Journal of Cancer. 2005 Sep;93(5):575–581. Available from: http://www.nature.com/articles/6602728.

39 Guan P, Howell-Jones R, Li N, Bruni L, de Sanjosé S, Franceschi S, et al. Human papillomavirus types in 115,789 HPV-positive women: a meta-analysis from cervical infection to cancer. International Journal of Cancer. 2012 Nov;131(10):2349–2359.

40 National Institute for Cancer Epidemiology and Registration (NICER) Foundation. Cancer incidence - National statistics on cancer incidence. Zurich, Switzerland; 2018. Available from: http://www.nicer.org/en/statistics-atlas/cancer-incidence/.

41 de Sanjose S, Quint WG, Alemany L, Geraets DT, Klaustermeier JE, Lloveras B, et al. Human papillomavirus genotype attribution in invasive cervical cancer: a retrospective cross-sectional worldwide study. The Lancet Oncology. 2010 Nov;11(11):1048–1056.

42 Walboomers JMM, Jacobs MV, Manos MM, Bosch FX, Kummer JA, Shah KV, et al. Human papillomavirus is a necessary cause of invasive cervical cancer worldwide. The Journal of Pathology. 1999 Sep;189(1):12–19. Available from: http://doi.wiley.com/10.1002/%28SICI%291096-9896%28199909%29189%3A1%3C12%3A%3AAID-PATH431%3E3.0.CO%3B2-F.

43 Federal Statistical Office. La population de la Suisse 2016. Neuchâtel, Suisse; 2017. 349–1600.

44 Federal Office of Public Health. Durchimpfung von 2-, 8-und 16-Jährigen Kindern in der Schweiz, 1999–2017;.

45 Commission Assurance Qualité. Avis d’expert No 18, mis à jour Janvier 2011 - Vaccination HPV. Bern, Schweiz: Gynécologie Suisse; 2011. 18.

46 Gold MR, Franks P, McCoy KI, Fryback DG. Toward Consistency in Cost-Utility Analyses: Using National Measures to Create Condition-Specific Values. Medical Care. 1998;36(6):778–792. Available from: https://www.jstor.org/stable/3766996.

47 Stratton KR, Durch J, Lawrence RS. Vaccines for the 21st century: a tool for decision-making. Washington, D.C.: National Academy Press; 2000.

48 Mandelblatt JS, Lawrence WF, Womack SM, Jacobson D, Yi B, Hwang Yt, et al. Benefits and Costs of Using HPV Testing to Screen for Cervical Cancer. JAMA. 2002 May;287(18):2372–2381. Available from: https://jamanetwork.com/journals/jama/fullarticle/194902.

49 Kim JJ, Goldie SJ. Health and Economic Implications of HPV Vaccination in the United States. New England Journal of Medicine. 2008 Aug;359(8):821–832. Available from: https://doi.org/10.1056/NEJMsa0707052.

50 World Health Organization. Making choices in health : WHO guide to cost-effectiveness analysis; 2003. Available from: http://apps.who.int/iris/handle/10665/42699.

51 Ultsch B, Damm O, Beutels P, Bilcke J, Brüggenjürgen B, Gerber-Grote A, et al. Methods for Health Economic Evaluation of Vaccines and Immunization Decision Frameworks: A Consensus Framework from a European Vaccine Economics Community. Pharmacoeconomics. 2016;34:227–244. Available from: https://www.ncbi.nlm.nih.gov/pmc/articles/PMC4766233/.

52 Damm O, Horn J, Mikolajczyk RT, Kretzschmar MEE, Kaufmann AM, Delere Y, et al. Cost-effectiveness of human papillomavirus vaccination in Germany. Cost Effectiveness and Resource Allocation. 2017 Sep;15:18. WOS:000409051400001.

53 Chesson HW, Markowitz LE, Hariri S, Ekwueme DU, Saraiya M. The impact and cost-effectiveness of nonavalent HPV vaccination in the United States: Estimates from a simplified transmission model. Human Vaccines & Immunotherapeutics. 2016;12(6):1363–1372.

54 Federal Statistical Office. Produit intérieur brut par habitant; 2018.

55 Marseille E, Larson B, Kazi DS, Kahn JG, Rosen S. Thresholds for the cost–effectiveness of interventions: alternative approaches. Bulletin of the World Health Organization. 2015 Feb;93(2):118–124. Available from: http://www.who.int/entity/bulletin/volumes/93/2/14-138206.pdf.

56 Schweizerische Konferenz der kantonalen Gesundheitsdirektorinnen und –direktoren (GDK), Conférence suisse des directrices et directeurs cantonaux de la santé (CDS), Conferenza svizzera delle direttrici e dei direttori cantonali della sanità. Die Impfung der Mädchen gegen Gebärmutterhalskrebs kann beginnen. Medienmitteilung; 2008.

57 Wolff E, Elfström KM, Haugen Cange H, Larsson S, Englund H, Sparén P, et al. Cost-effectiveness of sex-neutral HPV-vaccination in Sweden, accounting for herd-immunity and sexual behaviour. Vaccine. 2018 Aug;36(34):5160–5165. Available from: http://www.sciencedirect.com/science/article/pii/S0264410X18309484.

58 Tay SK, Hsu TY, Pavelyev A, Walia A, Kulkarni AS. Clinical and economic impact of school-based nonavalent human papillomavirus vaccine on women in Singapore: a transmission dynamic mathematical model analysis. BJOG: An International Journal of Obstetrics & Gynaecology. 2018 Mar;125(4):478–486. Available from: https://obgyn.onlinelibrary.wiley.com/doi/abs/10.1111/1471-0528.15106.

59 Mennini FS, Bonanni P, Bianic F, de Waure C, Baio G, Plazzotta G, et al. Cost-effectiveness analysis of the nine-valent HPV vaccine in Italy. Cost Effectiveness and Resource Allocation. 2017 Jul;15:11. WOS:000405203200001.

60 Hartwig S, St Guily JL, Dominiak-Felden G, Alemany L, de Sanjosé S. Estimation of the overall burden of cancers, precancerous lesions, and genital warts attributable to 9-valent HPV vaccine types in women and men in Europe. Infectious Agents and Cancer. 2017 Apr;12. Available from: https://www.ncbi.nlm.nih.gov/pmc/articles/PMC5387299/.

61 Simms KT, Laprise JF, Smith MA, Lew JB, Caruana M, Brisson M, et al. Cost-effectiveness of the next generation nonavalent human papillomavirus vaccine in the context of primary human papillomavirus screening in Australia: a comparative modelling analysis. The Lancet Public Health. 2016 Dec;1(2):e66–e75. Available from: http://www.sciencedirect.com/science/article/pii/S2468266716300196.

62 Kind AB, Pavelyev A, Mouaddin NE, Schmidt A, E Morais PG, Lienert F. Conference abstract: Epidemiologic impact of a gender-neutral nonavalent HPV vaccination programme in comparison to the current gender-neutral quadrivalent HPV vaccination programme in Switzerland. EUROGIN 2018;.

63 Brisson M, Edmunds WJ. Economic Evaluation of Vaccination Programs: The Impact of Herd-Immunity. Medical Decision Making. 2003 Jan;23(1):76–82. Available from: https://doi.org/10.1177/0272989X02239651.

64 Pitman R, Fisman D, Zaric GS, Postma M, Kretzschmar M, Edmunds J, et al. Dynamic Transmission Modeling: A Report of the ISPOR-SMDM Modeling Good Research Practices Task Force-5. Value in Health. 2012 Sep;15(6):828–834. Available from: http://www.sciencedirect.com/science/article/pii/S1098301512016518.

65 Laprise JF, Markowitz LE, Chesson HW, Drolet M, Brisson M. Comparison of 2-Dose and 3-Dose 9-Valent Human Papillomavirus Vaccine Schedules in the United States: A Cost-effectiveness Analysis. The Journal of Infectious Diseases. 2016 Sep;214(5):685–688. Available from: https://www.ncbi.nlm.nih.gov/pmc/articles/PMC4978371/.

66 Donken R, Bogaards JA, van der Klis FRM, Meijer CJLM, de Melker HE. An exploration of individual- and population-level impact of the 2-dose HPV vaccination schedule in pre-adolescent girls. Human Vaccines & Immunotherapeutics. 2016;12(6):1381–1393.

67 Harper DM, DeMars LR. HPV vaccines – A review of the first decade. Gynecologic Oncology. 2017 Jul;146(1):196–204. Available from: http://www.sciencedirect.com/science/article/pii/S0090825817307746.

68 International Agency for Research on Cancer. IARC monographs on the evaluation of carcinogenic risks to humans, volume 90, Human papillomaviruses. Lyon; 2007.

69 Gillison ML, Chaturvedi AK, Lowy DR. HPV prophylactic vaccines and the potential prevention of noncervical cancers in both men and women. Cancer. 2008 Nov;113(10):3036–3046. Available from: https://onlinelibrary.wiley.com/doi/abs/10.1002/cncr.23764.

70 Parkin DM. The global health burden of infection-associated cancers in the year 2002. International Journal of Cancer. 2006 Jun;118(12):3030–3044. Available from: https://onlinelibrary.wiley.com/doi/abs/10.1002/ijc.21731.

71 Adams M, Jasani B, Fiander A. Human papilloma virus (HPV) prophylactic vaccination: Challenges for public health and implications for screening. Vaccine. 2007 Apr;25(16):3007–3013. Available from: http://www.sciencedirect.com/science/article/pii/S0264410X07000369.

72 Thamsborg LH, Andersen B, Larsen LG, Christensen J, Johansen T, Hariri J, et al. Danish method study on cervical screening in women offered HPV vaccination as girls (Trial23): a study protocol. BMJ Open. 2018 May;8(5):e020294. Available from: https://bmjopen.bmj.com/content/8/5/e020294.

73 Smith MA, Gertig D, Hall M, Simms K, Lew JB, Malloy M, et al. Transitioning from cytology-based screening to HPV-based screening at longer intervals: implications for resource use. BMC Health Services Research. 2016 Apr;16(1):147. Available from: https://doi.org/10.1186/s12913-016-1375-9.

74 Simms KT, Smith MA, Lew JB, Kitchener HC, Castle PE, Canfell K. Will cervical screening remain cost-effective in women offered the next generation nonavalent HPV vaccine? Results for four developed countries. International Journal of Cancer. 2016 Dec;139(12):2771–2780. Available from: https://onlinelibrary.wiley.com/doi/abs/10.1002/ijc.30392.

75 Kim JJ, Burger EA, Sy S, Campos NG. Optimal Cervical Cancer Screening in Women Vaccinated Against Human Papillomavirus. JNCI Journal of the National Cancer Institute. 2016 Oct;109(2). Available from: https://www.ncbi.nlm.nih.gov/pmc/articles/PMC5068562/.

76 Commission Assurance Qualité. Avis d’expert No 50. Gynécologie Suisse; 2018.

77 Lew JB, Simms KT, Smith MA, Hall M, Kang YJ, Xu XM, et al. Primary HPV testing versus cytology-based cervical screening in women in Australia vaccinated for HPV and unvaccinated: effectiveness and economic assessment for the National Cervical Screening Program. The Lancet Public Health. 2017;2(2):e96–e107.

